# *ZCWPW1* loss-of-function variants in Alzheimer’s Disease

**DOI:** 10.1101/2021.08.13.21261426

**Authors:** Fahri Küçükali, Katrin Nußbaumer, Jasper Van Dongen, Elisabeth Hens, Céline Bellenguez, Benjamin Grenier-Boley, Delphine Daian, Anne Boland, Jean-François Deleuze, Jean-Charles Lambert, Christine Van Broeckhoven, Kristel Sleegers, on behalf of the BELNEU Consortium

**Affiliations:** Complex Genetics of Alzheimer’s Disease Group, Center for Molecular Neurology, VIB, Antwerp, Belgium; Department of Biomedical Sciences, University of Antwerp, Antwerp, Belgium; University of Applied Sciences Campus Vienna, Vienna, Austria; Neurodegenerative Brain Diseases Group, VIB Center for Molecular Neurology, Antwerp, Belgium; Department of Neurology and Memory Clinic, Hospital Network Antwerp, Middelheim and Hoge Beuken, Antwerp, Belgium; Department of Neurology, University Hospital Antwerp, Edegem, Belgium; Department of Neurology, University Hospital Brussel and Center for Neurosciences, Free University of Brussels, Brussels, Belgium; Univ. Lille, Inserm, CHU Lille, Institut Pasteur de Lille, U1167-RID-AGE facteurs de risque et déterminants moléculaires des maladies liés au vieillissement, Lille, France; Université Paris-Saclay, CEA, Centre National de Recherche en Génomique Humaine, 91057, Evry, France

## Abstract

Genome-wide association studies (GWAS) have identified more than 75 genetic risk loci for Alzheimer’s Disease (AD), however for a substantial portion of these loci the genetic variants or genes directly involved in AD risk remain to be found. A GWAS locus defined by the index SNP rs1476679 in *ZCWPW1* is one of the largest AD loci as the association signal spans 56 potential risk genes. The three most compelling candidate genes in this locus are *ZCWPW1, PILRA* and *PILRB*, based on genetic, transcriptomic, and proteomic evidence. We performed amplicon-based target enrichment and next-generation sequencing of the exons, exon-intron boundaries, and UTRs of *ZCWPW1, PILRA* and *PILRB* on an Illumina MiSeq platform in 1048 Flanders-Belgian late-onset AD patients and 1037 matched healthy controls. Along with the single-marker association testing, the combined effect of Sanger-validated rare variants was evaluated in SKAT-O. No common variants (*n* = 40) were associated with AD. We identified 20 validated deleterious rare variants (MAF < 1%, CADD score ≥ 20), 14 of which in *ZCWPW1*. This included 4 predicted loss-of-function (LoF) mutations that were exclusively found in patients (*P* = 0.011). Haplotype sharing analysis revealed distant common ancestors for two LoF mutations. Single-molecule long-read Nanopore sequencing analysis unveiled that all LoF mutations are phased with the risk haplotype in the locus. Our results support the recent report for the role of ultra-rare LoF *ZCWPW1* variants in AD and suggest a potential risk mechanism for AD through *ZCWPW1* haploinsufficiency.

## Introduction

Alzheimer’s disease (AD) is a neurodegenerative brain disorder that is the most common cause of dementia^1^. AD is a complex disease with a high heritability estimate^2^; over the past decade more than 75 genetic susceptibility loci have been identified in genome-wide association studies (GWAS)^3–10^. One of these risk loci is denoted as the Zinc finger CW-type and PWWP domain containing 1 (*ZCWPW1*) locus, because the discovery lead SNP rs1476679 was located intronic in *ZCWPW1*^3^. The *ZCWPW1* locus is among the 10 most significant genetic risk loci for AD, but is also one of the most complex loci, as the association signal spans 620 Kb, comprising 56 genes of which 29 are protein-coding, including nearby genes harboring other genome-wide significant SNPs, such as the Paired Immunoglobulin-Like Type 2 Receptor Alpha (*PILRA*) and Beta (*PILRB*) genes (Figure 1). Four variants across the locus were reported as lead variant with similar effect size and direction in seven subsequent large-scale AD GWAS: (i) the discovery lead SNP rs1476679^4^, (ii) exonic common variant rs1859788^5,7,8^ in *PILRA* (p.Gly78Arg), (iii) 3’ UTR variant rs12539172^6^ in Neuronal Tyrosine Phosphorylated Phosphoinositide-3-Kinase Adaptor 1 (*NYAP1*), and (iv) intronic variant rs7384878^9,10^ in *PMS2P1* (PMS1 Homolog 2, Mismatch Repair System Component Pseudogene 1) which is a pseudogene located between *SPDYE3* (Speedy/RINGO Cell Cycle Regulator Family Member E3) and *PILRB* (Figure 1). This phenomenon, caused by linkage disequilibrium (LD) and relatively high gene density in the locus, complicates the interpretation of this association signal in terms of mechanism of action in Alzheimer’s disease. Subsequent whole exome and whole genome sequencing studies, expression quantitative trait loci (eQTL) based studies, and functional studies have proposed several different candidate genes in the locus.

**Figure 1.**
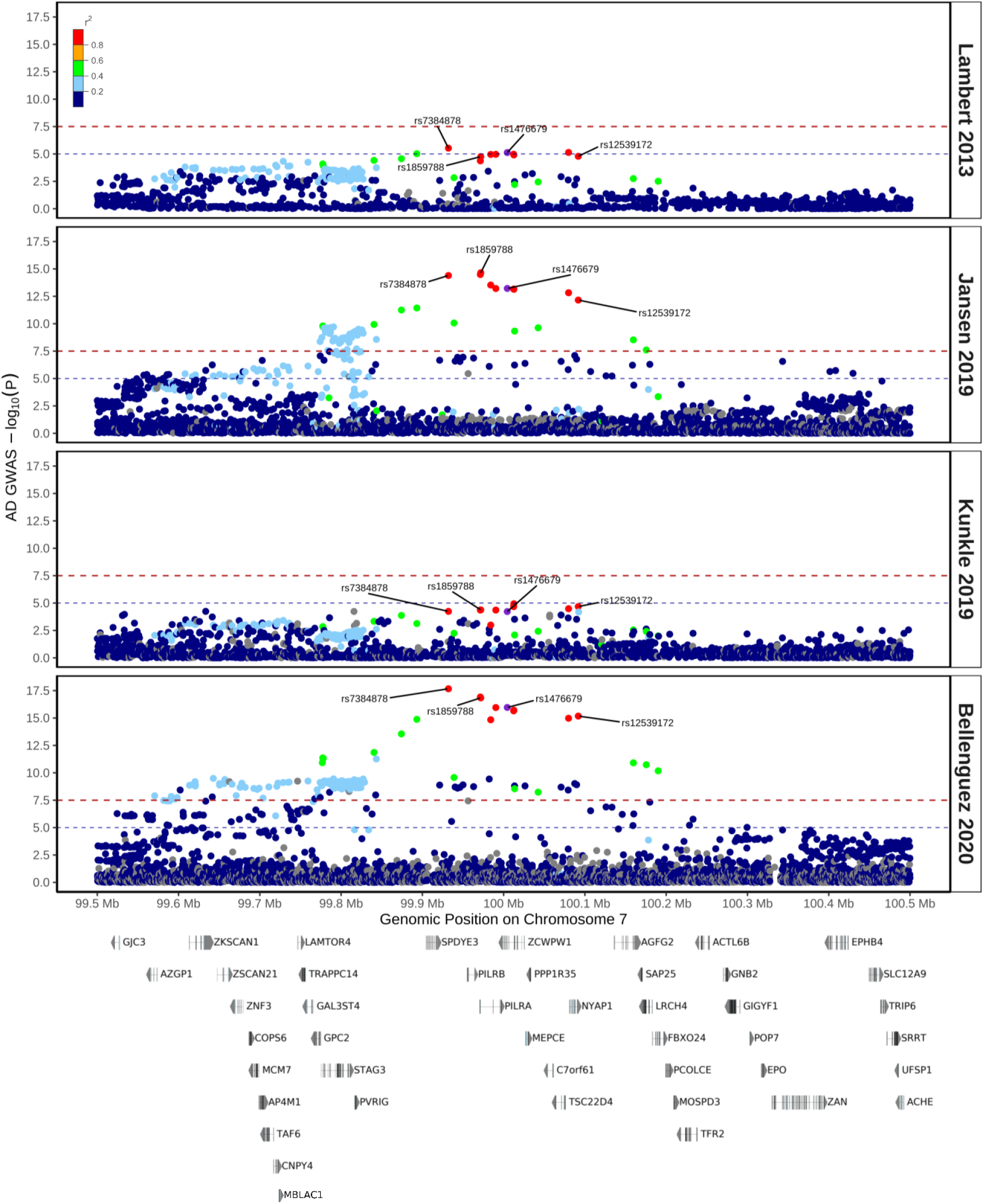
AD GWAS association signals in the *ZCWPW1* locus. The comparison of the regional plots for the AD association signals in four main GWAS which each first described one of the four main lead variants currently known in the literature. The discovery stage (Stage 1) results of the variants between hg19 chr7:99.5-100.5 Mb coordinates were used for plotting the association signals in each GWAS. The linkage disequilibrium r^2^ values were calculated with respect to the discovery GWAS index variant rs1476679 (marked in purple) in the 1KG EUR ancestry samples. The four lead variants were annotated on the regional plot. The blue dashed line represents suggestive significant (*P* ≤ 1E-5) threshold, meanwhile the red dashed line represents the genome-wide significance threshold (*P* ≤ 5E-8). The relative positions of the canonical transcripts of the protein-coding genes in the locus (based on GENCODE v38) were described in the bottom part of this figure.

First, a UK whole exome sequencing (WES) project observed an increased burden of *PILRA* variants in AD patients as their top-associated hit, suggesting a direct role of *PILRA* in AD pathogenesis^11^. In contrast, one of the largest WES studies on AD reported to date pinpointed significant enrichment of ultra-rare (allele frequency < 0.01%) *ZCWPW1* predicted loss-of-function (LoF) variants in AD patients^12^, and additionally, a nominally significant association of genic rare (allele frequency < 1%) variants within *ZCWPW1* was reported in a recent large-scale WGS study on AD^13^. Second, the discovery lead SNP in the locus, rs1476679, has strong regulatory potential, with the top RegulomeDB^14^ rank score among the lead SNPs of the significant loci from all published AD GWAS, and two independent studies observed that rs1476679 is an eQTL for *PILRB*^15,16^. However, annotation with other eQTL catalogues (GTEx^17^, Brain xQTL Serve^18^, eQTLGen^19^, and numerous datasets analyzed uniformly by eQTL Catalogue database^20^) of AD-relevant tissues and cell types reveals that rs1476679 is also a significant eQTL for other genes in the locus, including *ZCWPW1* and *PILRA*. Furthermore, the GWAS association signal in the locus was found to be highly colocalized with lipopolysaccharide-stimulated monocyte eQTL signals for *ZCWPW1* and *PILRA*^8^. Moreover, it was suggested that rs1476679 might be controlling the expression of nearby genes via CTCF-mediated chromatin loops^21^. Third, a ligand binding assay based functional study proposes that the lead SNP rs1859788, encoding the amino acid substitution p.Gly78Arg in *PILRA*, might be a protective variant in the locus as it potentially reduces inhibitory signaling in microglia^22^.

The literature on the function of PILRB and PILRA proteins indicates that they operate as activating and inhibitory receptors (respectively) for the regulation of the immune response. PILRA contains an immunoreceptor tyrosine-based inhibitory motif (ITIM) in its cytoplasmic tail that recruits phosphatases such as PTPN6/SHP-1, meanwhile PILRB is thought to act through pairing with immunoreceptor tyrosine-based activation motif (ITAM)-containing proteins such as TYROBP/DAP12^23^. On the other hand, until recently, the function of ZCWPW1 protein was largely unknown, and a role in epigenetic regulation as a histone modification reader (especially for H3K4me3) was proposed based on the structure and biochemical information of its zf-CW domain^24^. Recently, ZCWPW1 function was studied for the first time in consecutive studies which showed that it is required during meiosis prophase I in a sex-dependent manner in mice^25^ and it is essential for repairing of double strand breaks (DSBs) marked by histone modification writer PRDM9 during meiosis^26–28^.

Taken together, it is not clear yet how this locus contributes to AD; however, the current literature prioritizes three genes in the locus: *ZCWPW1, PILRA* and *PILRB*. To this end, in this study we aimed to investigate genetic variation in the locus by sequencing UTRs, exons and exon-intron boundaries of these three genes in our well-characterized case-control cohort.

## Materials and Methods

### Study Cohort

The study cohort consists of 2085 Flanders-Belgian individuals. Patients were ascertained at the Memory and Neurology Clinics of the BELNEU consortium. The patient group consisted of 1048 late-onset AD cases, of whom 65.4% female, with a mean onset age of 78.8 ± 5.7 years. Possible, probable, and definite AD diagnosis was based on NINCDS-ADRA^29^ and/or NIA-AA^30,31^ criteria. Control individuals were recruited from partners of patients, or were volunteers from the Belgian community, and included 1037 age-matched healthy individuals (64.8% female and mean inclusion age of 70.2 ± 9.1 years). All control individuals scored >25 on the Montreal Cognitive Assessment (MoCA)^32^ test, and were negative for subjective memory complaints, neurological or psychiatric antecedents, and family history of neurodegeneration. Both patient and control group individuals originated from the same geographical area (Flanders-Belgium), with no evidence of population stratification (Supplementary Figure 1). All participants and/or their legal guardian signed written informed consent forms before inclusion. The study protocols were approved by the ethics committees of the Antwerp University Hospital and the University of Antwerp, and the ethics committees of the participating neurological centers of the BELNEU consortium.

### Targeted Resequencing

For sequencing of regions of interest, we used genomic DNA (gDNA) samples that were amplified using Illustra GenomiPhi v2 kit (Thermo Fisher, Waltham, MA, USA). We targeted UTRs, exon-intron boundaries (20 bp long) and exons of all protein-coding transcripts of *PILRB, PILRA* and *ZCWPW1* genes based on GENCODE v19. Total length of targeted regions was 9185 bp. A custom-designed multiplex target enrichment assay (Agilent, Santa Clara, CA, USA) was prepared and optimized prior to sequencing. We amplified 47 amplicons targeting 43 exons of interest using primers with universal adapter sequences (primer sequences available upon request). Prepared amplicon libraries were then barcoded for each sample with Nextera XT sequences (Illumina, San Diego, CA, USA) targeting previously incorporated adapters. Indexed libraries were 2 × 300 bp paired-end (MiSeq Reagent Kit v3) sequenced on a MiSeq platform (Illumina, San Diego, CA, USA) in a total of 5 runs. Two amplicons (*PILRB* exon 5 and *ZCWPW1* exon 14) that did not achieve sufficient coverage were additionally sequenced by Sanger sequencing, consisting of PCR amplification, followed by dideoxy-termination with BigDye termination cycle sequencing kit v3.1 (Thermo Fisher) and sequencing with ABI3730 DNA Analyzer (Thermo Fisher). Resulting chromatograms were analyzed independently and blindly by two investigators using novoSNP^33^ and Seqman (DNAstar, Madison, WI, USA). All deleterious rare variants (CADD score ≥ 20 and MAF < 1%) and other rare variants of interest were validated using Sanger sequencing on gDNA of all carriers, as described above.

### Bioinformatic Processing

Adapter clipping of demultiplexed FASTQs was performed with fastq-mcf. Reads were aligned to the hg19 reference genome with Burrows-Wheeler Aligner MEMv0.7.5^34^. We merged duplicate sample aligned reads belonging to the same individual using SAMtools^35^ (v1.6) merge. Sequencing coverage was analyzed using SAMtools depth and visualized with Circos^36^ plot (Supplementary Figure 2).

Variants were called with GATK 4.1.4^37^ HaplotypeCaller. Multiallelic variant sites were split into biallelic records and raw indels were left-aligned and normalized using BCFtools (v1.9) norm. Variants were annotated with dbSNP build 151. Variant quality control was done by using standard hard filtering parameters according to the GATK Best Practices^38,39^. Additionally, with VCFTools^40^ (v0.1.16), we applied genotype level QC by assigning individual genotypes with genotype quality (GQ) < 20 and depth (DP) < 8 as missing. Moreover, using vcffilterjdk7^41^ with a custom filtering java code (available upon request), we also assigned genotypes as missing if heterozygous genotypes have allele depth ratios exceeding 1:3 ratio, and if homozygous genotypes do not have allele depth ratios larger than 9:1 or smaller than 1:9 ratios. Finally, we removed variants that were missing in more than 20% of the study cohort, deviated from Hardy-Weinberg Equilibrium (HWE *P* < 10^−6^ in controls) or differentially missing between cases and controls (*P* < 10^−6^). Variants were annotated with ANNOVAR^42^, SnpEff^43^ and CADD v1.6^44^. We used the latest Ensembl release (104) to select the canonical transcripts of *PILRB* (ENST00000609309.2), *PILRA* (ENST00000198536.7), and *ZCWPW1* (ENST00000684423.1), which were used as reference transcripts in this study to report the predicted functional consequences of the identified variants. Variant frequencies were queried in the most recent versions of the Genome Aggregation Database^45^ (gnomAD, v2.1.1), the Exome Aggregation Consortium^46^ (ExAC, v1) and Healthy Exomes^47^ (HEX, v1) browsers.

### Genotyping Array Data

Genotyping array data were available for *n*=1803 samples included in this study through the framework of European DNA Biobank (EADB) project. All pre- and post-genotyping sample and variant quality control procedures were described previously^9^. These data were used for (i) investigating the genetic ancestry of the included samples to control for population stratification, (ii) refining possible haplotypes of predicted LoF mutation carriers, (iii) investigating pairwise identity by descent (IBD) estimates between predicted deleterious mutation carriers both in a genome-wide (for checking cryptic relatedness) and locus specific manner, and (iv) obtaining genotypes for intronic rs34919929 variant that is in LD with GWAS discovery lead SNP rs1476679 and more proximal to *ZCWPW1* LoF variants of interest for Nanopore sequencing experiments. Both genetic principal components and pairwise IBD estimates were calculated in PLINK on quality-controlled, common (MAF ≥ 1%), non-missing (variant missingness ≤ 0.02), and LD-pruned (PLINK parameters: “-- indep-pairwise 500kb 1 0.2”) variants that were out of the long-range LD loci that are likely to confound genomic scans: LCT (2q21), HLA (including MHC), 8p23 and 17q21.31 inversions, and 24 other long-range LD regions^48^. The genetic principal components were analyzed in a PCA plot together with the *n=*2504 individuals from the 1000 Genomes (1KG) Project^49^.

### Statistical Analyses

Single marker association testing of the variants in the targeted regions was performed in PLINK^50^ (v1.9), using a χ^2^ allelic test to compare allele frequencies of rare variants (MAF < 1%) between patients and controls, and a logistic regression model for common variants (MAF ≥ 1%) to investigate the additive effect of the minor allele on Alzheimer’s disease status, adjusted by sex and age at onset (for patients) or age at inclusion (for control individuals) covariates. Odds ratios are reported for the minor allele with 95% confidence intervals (CI). Statistical significance thresholds are adjusted for multiple testing by Bonferroni-correction.

For gene-based rare variant (MAF < 1%) association analysis, the rare variants were classified into two groups based on their predicted functional consequences on the canonical transcripts of the genes as protein-altering (i.e. missense, nonsense, frameshift, and splice site disrupting) and predicted LoF (i.e. nonsense, frameshift, and splice site disrupting) variants and collapsed per gene. Gene-based optimized sequence kernel association test (SKAT-O)^51^ was used (“SKATBinary” function with ‘method = “SKATO”’ option, available in R package SKAT v1.3.2.1) to assess the combined effect of these rare variants on Alzheimer’s disease.

### Haplotype Sharing Analysis

A short tandem repeat (STR) panel was designed to investigate the possible haplotypes shared by non-singleton *ZCWPW1* predicted LoF mutation carriers. We preselected STRs described in Marshfield catalogue^52^ based on the following criteria: (i) at least 70% heterozygosity, (ii) proximity to locus of interest up to 5 million base-pairs, (iii) non-overlapping STR with different sizes. Eleven Marshfield STRs following these criteria and spanning 8.9 Mb and 5.33 cM were selected; six upstream STRs: D7S2431, D7S1796, D7S554, D7S651, D7S647, D7S2480, and five downstream STRs: D7S477, D7S666, D7S2448, D7S2504, D7S2494. The primers targeting these STRs were designed to contain HEX or FAM fluorescent labels, and the reactions were run in two-plex format. STR lengths were scored independently and blindly by two investigators using LGV (v3.01) in-house software. A subset of Flanders-Belgian control individuals (*n =* 172) were also genotyped with the same STR panel to estimate the frequencies of STR lengths in the study population.

### Nanopore Sequencing

Single-molecule long-read gDNA sequencing was performed on a MinION platform (Oxford Nanopore Technologies (ONT), Oxford, UK) to investigate phase information of the risk haplotype with *ZCWPW1* predicted LoF mutations. We chose rs34919929 for phasing with predicted LoF variants in the locus as rs34919929 is in linkage disequilibrium with GWAS index SNP rs1476679 (r^2^=0.98 in European population) and more proximal to *ZCWPW1* LoF variants of interest, allowing the design of shorter amplicons. We designed primers (Supplementary Table 1) with ONT adapter using Primer3^53^ (v4.0.1) to generate two amplicons spanning rs34919929 and *ZCWPW1* LoF mutations. The first amplicon (∼2.2 Kb) contained sites for rs34919929 and LoF mutation rs774275324. The second amplicon (∼6.5 Kb) contained sites for rs34919929, and LoF mutations rs774275324 and rs1180932049. The long amplicon spanned the short amplicon, however we decided to include both amplicons in the experiment to make use of the relatively better amplification efficacy of the short amplicon. Two amplicons were generated for two rs774275324 carriers that are heterozygous for the risk haplotype, two rs1180932049 carriers that are heterozygous for the risk haplotype and one rs1180932049 carrier that is homozygous for risk haplotype (as positive control). Amplicons generated for another three control samples without LoF mutations and with homozygous reference, heterozygous and homozygous alternative genotypes for rs34919929 variant were included in the experiment as negative controls. All PCR amplifications were performed with 35 cycles using KAPA LongRange HotStart PCR Kit KK3502 (KAPA Biosystems, MA, USA). After the reactions, excess primers and nucleotides were cleaned with ExoSAP-IT (Thermo Fisher).

Amplicons then were barcoded with the PCR Barcoding Expansion 1-96 kit (Oxford Nanopore Technologies) using 20 amplification cycles on a 1/200 diluted template. After purification with Agencourt AMPure XP beads (Beckman Coulter, Brea, CA, USA) and concentration measurement with Qubit (Thermo Fisher), amplicons were pooled equimolarly. The sequencing library was prepared as previously described^54^. SQK-LSK109 chemistry and FLO-MIN106 flow cell were used for sequencing.

Base calling of the raw reads was performed with ONT basecaller Guppy (v3.0.3) on the Promethion compute device. After demultiplexing of basecalled FASTQ reads with qcat (v1.0.1), alignment of demultiplexed reads to hg19 reference genome was performed with minimap2^55^ (v2.17). Variants were called with bcftools mpileup. Aligned reads were visualized in Integrative Genomics Viewer^56^ (IGV; v2.8.0) and phasing of variants of interest was performed using WhatsHap^57^ (v0.18).

## Results

### Single Marker Association

A total of 292 quality-controlled variants were identified in the targeted regions, of which 40 were common (MAF ≥ 1%). Chi-squared allelic association and logistic regression tests showed no association of any variants after Bonferroni correction. The GWAS index variants in the locus did show similar size and direction of effects to the original GWAS reports (Supplementary Table 2).

Four rare variants were nominally associated with Alzheimer’s disease (Supplementary Table 3). Of those, rs531189257 was exclusively found in 10 patients (MAF in AD patients = 0.53%). rs531189257 is a rare (gnomAD NFE MAF = 0.44%) intronic variant in the canonical *PILRA* transcript, and a 3’ UTR variant on the short transcript of *PILRA* (ENST00000484934.1).

### Deleterious Rare Variants and Gene-based Rare Variant Association

We then sought to investigate the rare variants (MAF < 1%) that are predicted to be deleterious (with either CADD ≥ 20 or “high impact” annotation by SnpEff), which includes mostly predicted LoF variants and high impact missense variants. We identified 20 validated deleterious variants (Table 1 and Figure 2).

**Table 1.**
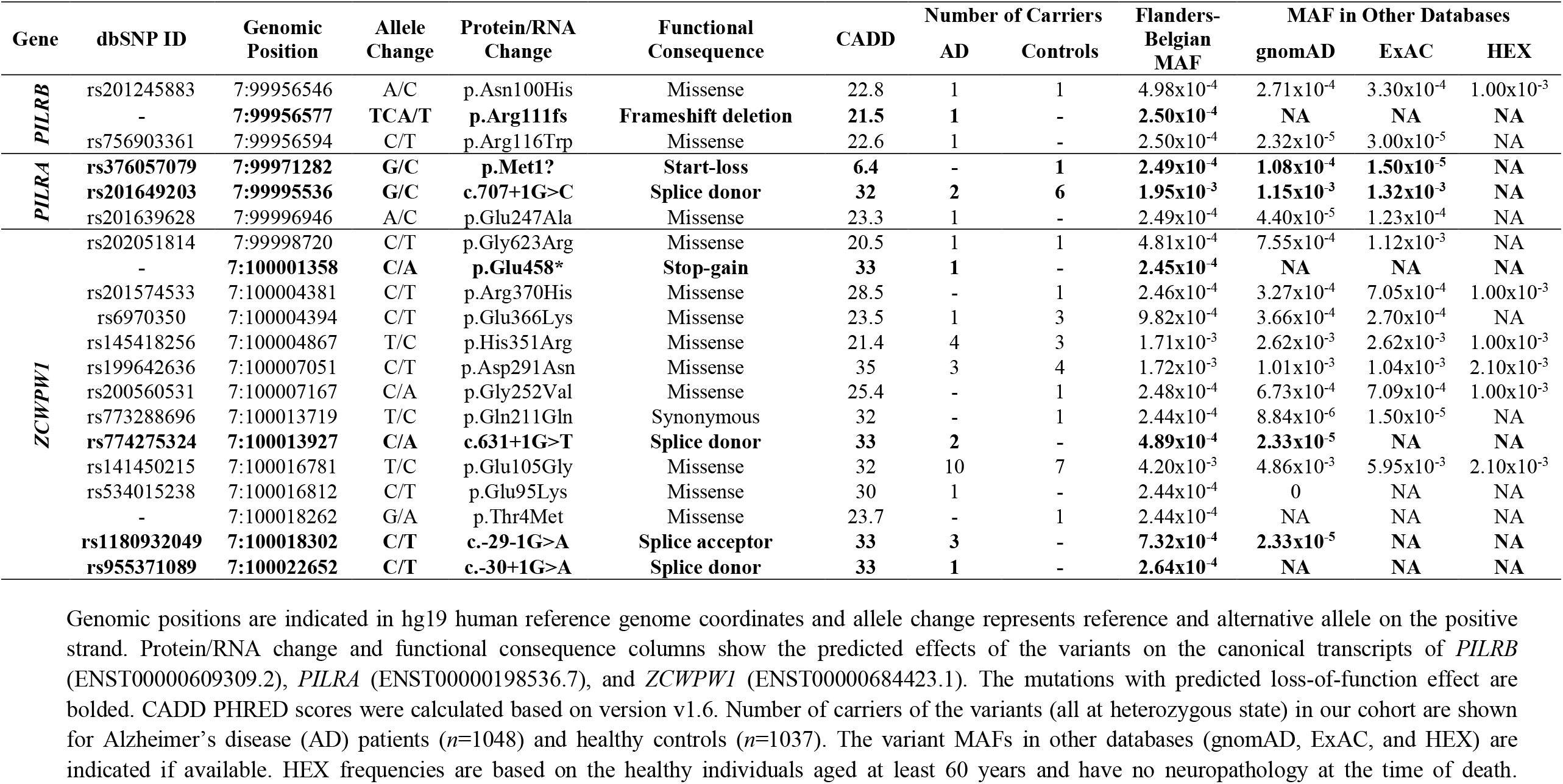
Rare deleterious variants in the targeted regions of *PILRB, PILRA* and *ZCWPW1*.

**Figure 2.**
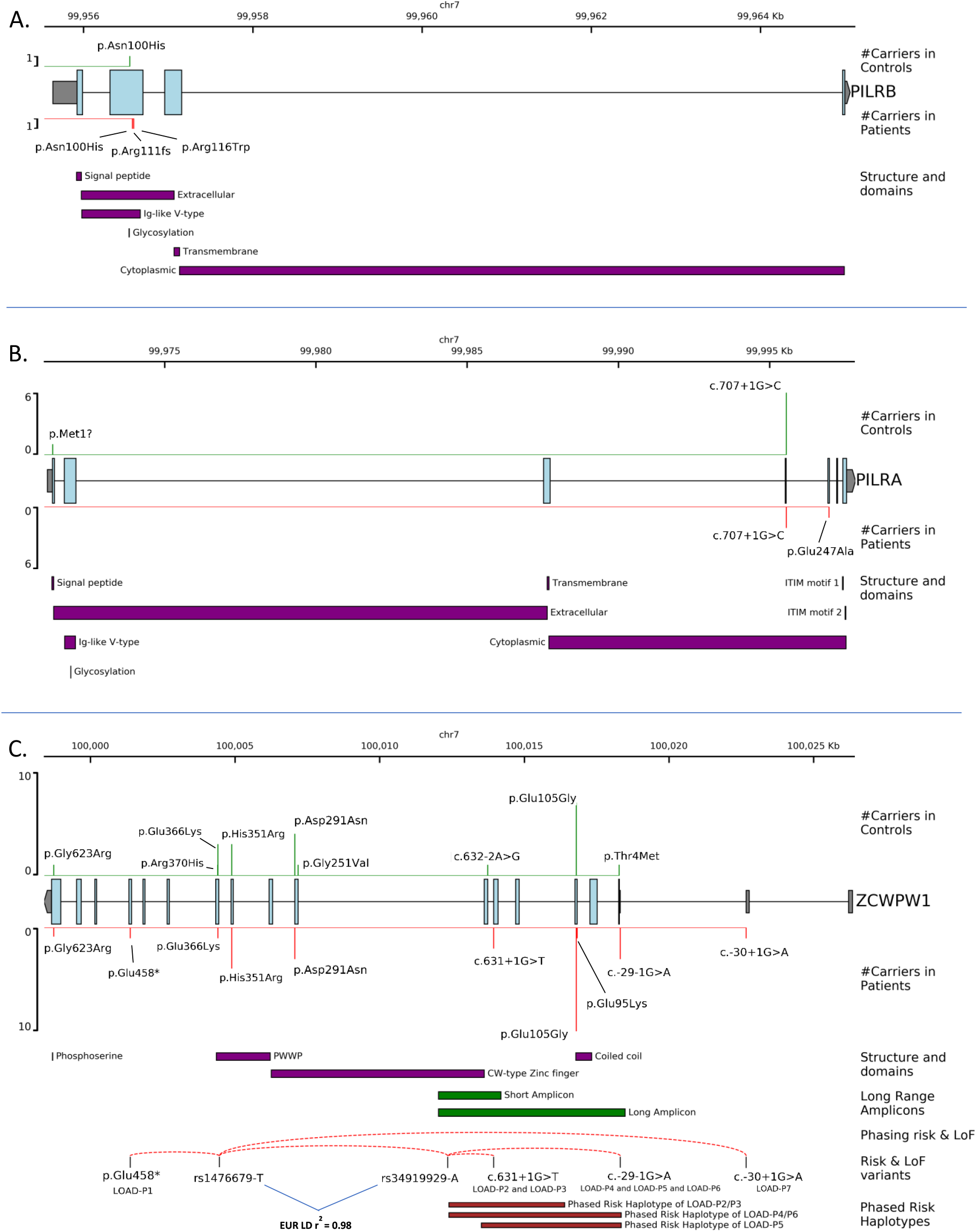
The schematic representation of the identified predicted deleterious variants. **(A)** The predicted deleterious variants identified in *PILRB*. **(B)** The predicted deleterious variants identified in *PILRA*. **(C)** The predicted deleterious variants identified in *ZCWPW1* along with the representation of the results of single molecule phasing of *ZCWPW1* predicted LoF variants. For each gene, the canonical transcripts (ENST00000609309.2, ENST00000198536.7, and ENST00000684423.1 respectively for *PILRB, PILRA*, and *ZCWPW1*) were plotted; with the light blue color indicates protein-coding sequences and the gray color indicates non-coding (UTR) sequences of the transcript. The relative positions of the identified predicted deleterious variants are shown on these transcripts. In order, the track descriptions are: the number of mutation carriers in healthy controls (*n*=1037; shown in green) and AD patients (*n*=1048; shown in red), known structures, motifs, posttranslational modification sites, topological domains, and functional domains of canonical protein isoforms retrieved from UniProt website, shown in purple. Additionally in **(C)** where patient-only *ZCWPW1* LoF variants were identified, the extra tracks represent (in order): long range amplicons designed for single molecule phasing experiment, the outcome of phasing experiments shown in red dashed archs connecting risk alleles with the LoF mutated alleles, the risk variants and LoF mutations of interest (and carrier IDs of those), and finally the coordinates of the risk haplotype for each LoF carriers spanning proxy risk allele rs34919929-C and LoF mutated alleles, as phased by WhatsHap (details were shown in Supplementary Table 5 and Supplementary Figures 5-7).

Three deleterious rare variants are located in *PILRB*, three in *PILRA* and fourteen in *ZCWPW1*. These include one novel frameshift mutation (p.Arg111fs) in *PILRB*, one novel stop-gain (p.Glu458*) and one novel missense (p.Thr4Met) mutation in *ZCWPW1*. All deleterious variants have a CADD score above 20, with the exception for *PILRA* p.Met1? start-loss mutation. As a result of the *PILRB* p.Arg111fs frameshift mutation (identified in a patient), a premature stop codon is introduced on the last exon of *PILRB* (at amino acid position 219), thus nonsense mediated decay (NMD) is not expected for this transcript. However, *PILRB* function of the mutated product is most likely impaired due to loss of nearly half of the canonical amino acid sequence that translates into topological and functional domains. *PILRA* seemed to be more tolerant against predicted LoF variants, as both variants were observed more in controls than in patients. On the other hand, we observed that predicted LoF variants (*n*=4) in *ZCWPW1* were exclusive to seven patients. These validated ultra-rare variants (gnomAD MAF < 0.01%) consisted of one novel stop-gain mutation and three splice site mutations. The stop-gain mutation, p.Glu458* that occurs at the 458^th^ amino acid of the canonical ZCWPW1 isoform whose total length is 649 amino acids, is predicted to result in haploinsufficiency through NMD. Furthermore, we identified two splice donor mutations (*ZCWPW1* c.631+1G>T and c.-30+1G>A) and one splice acceptor mutation (*ZCWPW1* c.-29-1G>A) in five patients. Interestingly, one predicted LoF mutation carrier (*ZCWPW1* c.-30+1G>A) also carried two predicted deleterious rare missense mutations (*ZCWPW1* p.Glu105Gly and *ZCWPW1* p.Glu95Lys, occurring *in cis* according to IGV investigation of sequencing reads as shown in the Supplementary Figure 4) within the coiled coil structure motif of the protein.

We investigated the combined effect of rare variants per gene using two classes of mutation: protein-altering and predicted LoF, revealing significant (*P* = 0.011) enrichment for *ZCWPW1* predicted LoF rare variants (Table 2). We did not observe any significant associations for protein-altering variants with Alzheimer’s disease in any of the genes.

**Table 2.**
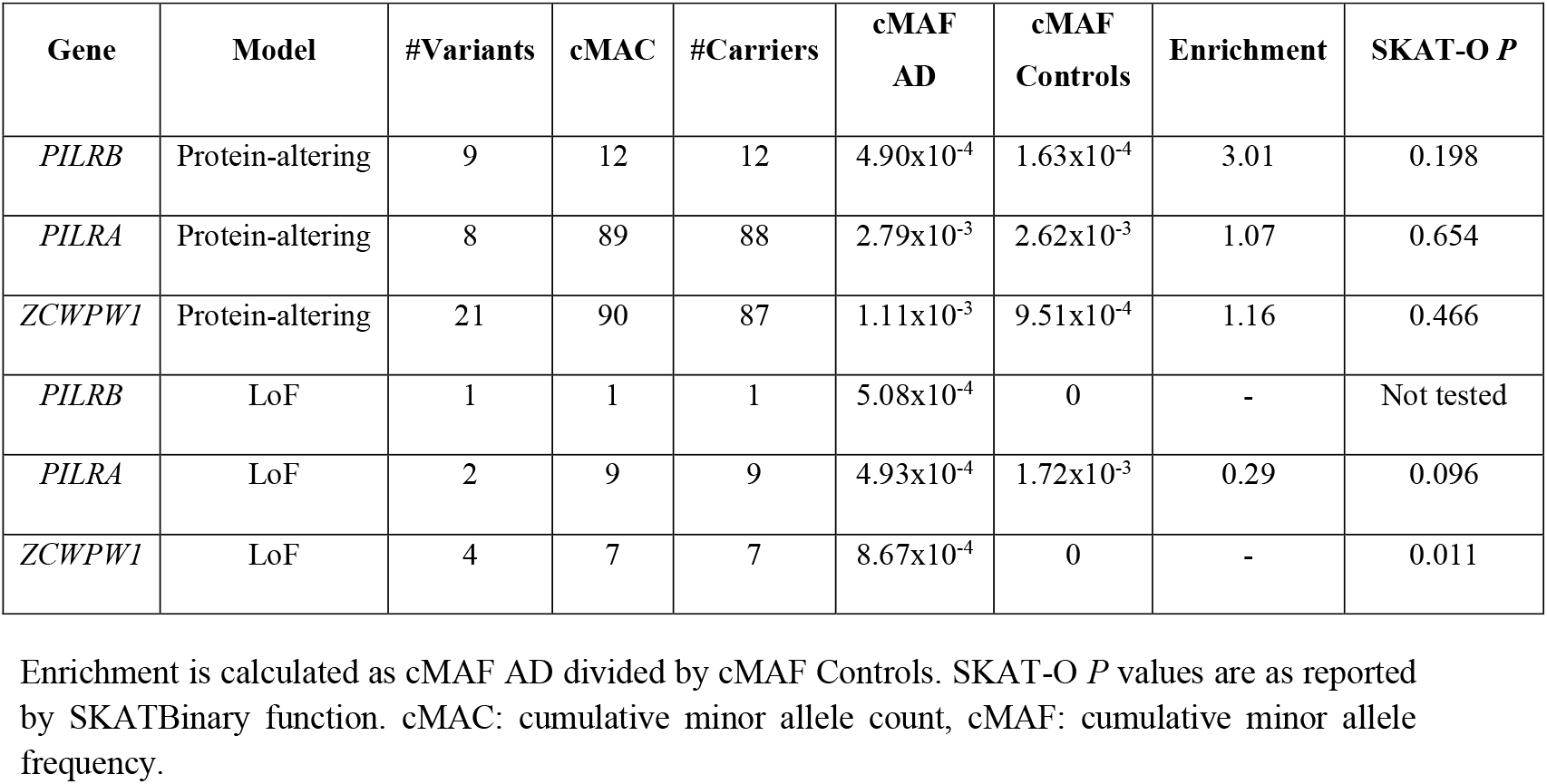
Gene-based SKAT-O results for rare variants per gene and models.

*ZCWPW1* predicted LoF carriers were all clinically diagnosed with probable or possible AD. Only carriers of the c.-29-1G>A mutation were diagnosed with additional features (frontal deficits (*n*=2), evidence of a vascular component (*n*=1)). The onset ages were rather high (mean onset age 77.7 ± 5.87 years), with exception of one carrier (69 years) who was also *APOE* ε4 homozygous. Otherwise, we did not observe a correlation between onset age and *APOE* genotype. Two of the three *ZCWPW1* c.-29-1G>A carriers had a familial history of the disease in the first degree. We did not have the biomaterials of these relatives to explore possible segregation of the mutation.

### Haplotype Sharing Analysis

Because we observed two of these ultra-rare predicted LoF variants in multiple patients, we investigated allele sharing in the *ZCWPW1* susceptibility locus in these carriers (LOAD-P2 and LOAD P3 for *ZCWPW1* c.631+1G>T and LOAD-P4, LOAD-P5 & LOAD-P6 for *ZCWPW1* c.-29-1G>A, see Supplementary Figure 3).

STR-based haplotype analyses revealed that *ZCWPW1* c.631+1G>T carriers shared alleles at eight consecutive STR markers, spanning a region of 5.5 to 8.3 Mb. The three carriers of *ZCWPW1* c.-29-1G>A shared alleles at six consecutive STR markers spanning a region of 4 to 5.5 Mb. Frequencies of these STR lengths are varying between 18.8% and 34% in the Flanders-Belgian population. LOAD-P4 and LOAD-P5 shared alleles at three additional distal markers, LOAD-P5 and LOAD-P6 shared alleles at three additional distal markers, and LOAD-P4 and LOAD-P6 shared additional alleles at five distal markers. Moreover, we used available genotyping array data of *ZCWPW1* predicted LoF carriers (excluding LOAD-P5 for whom these data were not available) to further refine these possible haplotypes constructed by STR markers. Comparison of *n*=1789 quality-controlled common variants between D7S2431 and D7S2494 revealed that the common haplotype between *ZCWPW1* c.631+1G>T carriers is limited by rs6970990 upstream and rs727708 downstream (the most proximal segments where identity by state [IBS] is not observed), spanning a region of 3.5 to 5.4 Mb. Similarly, the common SNPs rs1488514 and rs10245317 adjusted possible common haplotype length between LOAD-P4 and LOAD-P6 to a region of 6.9 to 7.9 Mb. All common SNPs in the highlighted haplotypes (Supplementary Figure 3) were shared by *ZCWPW1* predicted LoF carriers, supporting STR genotyping results.

Furthermore, we investigated pairwise identity-by-descent (IBD) between six *ZCWPW1* predicted LoF carriers based on whole-genome and locus-specific (limited to possible haplotypes shown in Supplementary Figure 3) SNP genotype data. Genome-wide pairwise IBD estimates (Supplementary Table 4) confirmed that these carriers are not close relatives (the highest pairwise IBD estimate PI-HAT = 0.015 between LOAD-P4 and LOAD-P3). However, when only locus-specific LD-pruned high-quality common variants are considered, we found strongly increased IBD proportions for carriers of the same mutation (LOAD-P2 – LOAD-P3 PI-HAT = 0.65 and LOAD-P4 – LOAD-P6 PI-HAT = 0.5), indicative of ancestral DNA segments (Supplementary Table 4).

### Nanopore Sequencing

We further investigated the possible relationship between identified *ZCWPW1* predicted LoF variants and the risk haplotype in the locus defined by genome-wide associated variant(s) in the *ZCWPW1* locus. We observed that 3 out of 7 *ZCWPW1* LoF variant carriers (LOAD-P1, LOAD-P5, LOAD-P7) have homozygous genotype for the risk allele of the index variant rs1476679 (TT), therefore these LoF variants are on the risk haplotype. In order to assess the phasing of the risk haplotype (using the proxy SNP rs34919929) and the LoF variants of the other four carriers, we performed single-molecule long-read sequencing as shown in Figure 2.

WhatsHap phasing (Supplementary Table 5) of these single molecule long-reads revealed that all LoF variants are phased with the risk haplotype in *ZCWPW1* locus. We confirmed these results by viewing WhatsHap-processed haplotype-tagged aligned reads on IGV (Supplementary Figures 5-7).

## Discussion

*ZCWPW1, PILRA* and *PILRB* have all been proposed as functional candidate genes in the *ZCWPW1* risk locus for Alzheimer’s disease. Here we report a targeted-resequencing study covering the UTRs, exons and splice sites of *PILRB, PILRA* and *ZCWPW1* in an AD case-control cohort of Flanders-Belgian origin.

The most significant result of this study was identification of ultra-rare *ZCWPW1* predicted LoF variants that were exclusively found in patients. Despite their low expected allele frequency based on gnomAD non-Finnish European (NFE) genotype frequency data, *ZCWPW1* c.631+1G>T and *ZCWPW1* c.-29-1G>A mutations were strongly enriched in our patient cohort (ORs for increased frequency compared to gnomAD NFE frequency were calculated as 20.56 [95% CI = 1.71-179.74] and 30.84 [95% CI = 4.13-230.11] respectively). Increased allele sharing in the locus between carriers of these mutations is compatible with a shared common ancestor, indicative of a founder effect in the Flanders-Belgian population. Long range haplotype phasing further demonstrated that all predicted LoF variants were located on the *ZCWPW1* risk haplotype defined by rs1476679 risk-increasing allele T, although it should be noted that this is the more common allele (gnomAD NFE allele frequency 70%).

Our findings are in line with a recent report from one of the largest WES study to date^12^, reporting enrichment of ultra-rare *ZCWPW1* predicted LoF variants in patients compared to controls (3.42 times enriched in patients, *P =* 0.02). In fact, this was the second most significant result among 24 AD-implicated genes after enrichment of ultra-rare *SORL1* predicted LoF variants in patients.

The significant enrichment identified for the predicted *ZCWPW1* LoF mutations in our cohort is mainly driven by three splice site mutations. *ZCWPW1* c.631+1G>T mutation occurs at the splice donor site of *ZCWPW1* exon 7, and a potential skipping event of exon 7 would disrupt the open reading frame (ORF) and introduce a premature stop codon in exon 8 (canonical amino acid position 221), meanwhile potential intron retention of intron 7 would also introduce a premature stop codon, thus both scenarios would lead to NMD of the mutated transcript. The other two splice site variants are in the 5’ UTR of *ZCWPW1* and occur in the intron 2 acceptor site (c.-29-1G>A) and donor site (c.-30+1G>A). Skipping exon 3 would cause exclusion of canonical start codon of *ZCWPW1*, and the earliest downstream ATG site on the same ORF could only produce a truncated protein lacking the first 68 amino acids if translation efficiency is adequate. A possible intron retention effect of these mutations would introduce a GC-poor (40%) 4.3 Kb long fragment before the canonical start codon and significantly decreasing the GC content of 5’ UTR of *ZCWPW1* that is naturally at 60%. Meanwhile, skipping of exon 2 would cause the exclusion of 109 bases prior to the canonical start codon that could alter translational efficacy and alternative splicing regulation. Moreover, the LoF effects of such 5’ UTR splice variants were previously reported for neurodegenerative diseases^58,59^.

The function of ZCWPW1 protein is not well-known, except for its involvement in epigenetic regulation as a histone modification reader^24^ and for its the recently proposed role during meiosis^25–27^. *ZCWPW1* has a consistent expression in different brain regions including hippocampus^17^, however its exact function in brain is yet to be examined. The current literature points out that loss of ZCWPW1 might be contributing to AD through inadequate epigenetic regulation as epigenetic mechanisms are suggested to be playing an important role in AD^60^. Furthermore, even though the only known significant brain eQTL effect of rs1476679 for *ZCWPW1* (identified in putamen part of basal ganglia) suggests a *ZCWPW1*-decreasing effect of the rs1476679-C protective allele^17^, the same protective allele is also significantly associated with increased *ZCWPW1* levels in blood^19^ and in stimulated monocytes^20,61^ for which substantial colocalization with GWAS was also observed^8^. The observation of predicted LoF mutations in *ZCWPW1* in AD patients aligns well with these observations. A recent single-nuclei RNAseq study^62^ on parietal lobe of AD brains showed upregulation of *ZCWPW1* in oligodendrocytes and downregulation in neurons. This suggests that these cell types should be prioritized when further investigating *ZCWPW1* loss-of-function variants. Moreover, as ZCWPW1 is thought to be an epigenetic regulator, it could be interesting to compare epigenetic profiles of brain tissues of LoF carriers with non-carriers in the future; differentially regulated regions could hint at the targets of ZCWPW1.

We should note that even though we assessed the rare coding variants in the most compelling genes of the *ZCWPW1* locus based on literature evidence, we cannot rule out the possibility that the rare variants in other genes more distal to the GWAS index variants could be contributing to Alzheimer’s disease.

Furthermore, the strongest association we obtained from our single marker testing was observed for rs531189257 intronic variant in *PILRA* that was exclusive to 10 patients (*P* = 0.002). This rare variant is also a 3’ UTR variant in a noncoding short transcript of *PILRA* (ENST00000484934.1) and it could be speculated that its possible effect on AD risk might be through regulatory effects on *PILRA*. However, it is important to note that replication is needed to further comment on the possible effects of this rare variant.

In conclusion, in this study we assayed three genes in the *ZCWPW1* AD risk locus and found four *ZCWPW1* predicted LoF mutations that were exclusively and significantly found in seven LOAD patients. Further investigation on these carriers revealed possible shared haplotypes hinting at distant common ancestors of non-singleton LoF carriers, meanwhile long-read Nanopore sequencing analysis unveiled that all LoF mutations are phased with the risk haplotype in the locus. Taken together, our results suggest a potential risk mechanism for AD through *ZCWPW1* haploinsufficiency, which should be supported with functional experiments in the future. Identification of *ZCWPW1* LoF variants in AD patients may open up new opportunities to study downstream effects of these mutations in Alzheimer’s disease.

## Data Availability

To comply with EU law and participant privacy, individual-level clinical and genetic data cannot be shared publicly, however summary statistics for single marker genetic association and gene-based rare variant association analyses can be found in the manuscript and the supplementary tables.

## Web Sources

fastq-mcf, https://github.com/ExpressionAnalysis/ea-utils/blob/wiki/FastqMcf.md

Phase 3 VCFs of 1KG samples, http://ftp.1000genomes.ebi.ac.uk/vol1/ftp/release/20130502/

GENCODE, https://www.gencodegenes.org/

BCFtools, http://samtools.github.io/bcftools/bcftools.html

qcat, https://github.com/nanoporetech/qcat

RegulomeDB, http://www.regulomedb.org/

CADD, https://cadd.gs.washington.edu/

Ensembl, https://www.ensembl.org/index.html

GTEx, https://gtexportal.org/home/index.html

eQTL Catalogue database, https://www.ebi.ac.uk/eqtl/

gnomAD browser, https://gnomad.broadinstitute.org/

ExAC browser, http://exac.broadinstitute.org/

Healthy Exomes (HEX) browser, https://www.alzforum.org/exomes/hex

Single-nuclei Brain RNA-seq expression browser, http://ngi.pub/snuclRNA-seq/

UniProt, https://www.uniprot.org/

pyGenomeTracks, https://github.com/deeptools/pyGenomeTracks

ggplot2: https://ggplot2.tidyverse.org/

## Acknowledgements

The authors thank the personnel of the VIB Neuromics Support Facility of the VIB-UAntwerp Center for Molecular Neurology and the personnel of the Human Biobank of the VIB NBD group. The research was funded in part by the Alzheimer Research Foundation (SAO-FRA), the Research Foundation Flanders (FWO), and the University of Antwerp Research Fund, Belgium. FK is a supported by a PhD fellowship from the University of Antwerp Research Fund. We thank the high-performance computing service of the University of Lille. We thank all the CEA-CNRGH staff who contributed to sample preparation and genotyping for GSA genotyping for their excellent technical assistance. This work was supported in part by a grant (European Alzheimer DNA BioBank, EADB) from the EU Joint Programme – Neurodegenerative Disease Research (JPND). Inserm UMR1167 is also funded by Inserm, Institut Pasteur de Lille, the Lille Métropole Communauté Urbaine, the French government’s LABEX DISTALZ program (development of innovative strategies for a transdisciplinary approach to Alzheimer’s disease).

## References

1. Gaugler, J., James, B., Johnson, T., Scholz, K. & Weuve, J. 2016 Alzheimer’s disease facts and figures. Alzheimer’s Dement. 12, 459–509 (2016).

2. Gatz, M. et al. Role of genes and environments for explaining Alzheimer disease. Arch. Gen. Psychiatry 63, 168–174 (2006).

3. Lambert, J.-C. et al. Meta-analysis of 74,046 individuals identifies 11 new susceptibility loci for Alzheimer’s disease. Nat. Genet. 45, 1452–1458 (2013).

4. Marioni, R. E. et al. GWAS on family history of Alzheimer’s disease. Transl. Psychiatry 8, 8–14 (2018).

5. Jansen, I. E. et al. Genome-wide meta-analysis identifies new loci and functional pathways influencing Alzheimer’s disease risk. Nat. Genet. 51, 404–413 (2019).

6. Kunkle, B. W. et al. Genetic meta-analysis of diagnosed Alzheimer’s disease identifies new risk loci and implicates Aβ, tau, immunity and lipid processing. Nat. Genet. 51, 414–430 (2019).

7. de Rojas, I. et al. Common variants in Alzheimer’s disease and risk stratification by polygenic risk scores. Nat. Commun. 12, 3417 (2021).

8. Schwartzentruber, J. et al. Genome-wide meta-analysis, fine-mapping and integrative prioritization implicate new Alzheimer’s disease risk genes. Nat. Genet. (2021) doi:10.1038/s41588-020-00776-w.

9. Bellenguez, C. et al. New insights on the genetic etiology of Alzheimer’s and related dementia. medRxiv (2020) doi:https://doi.org/10.1101/2020.10.01.20200659.

10. Wightman, D. P. et al. Largest GWAS (N=1,126,563) of Alzheimer ‘s Disease Implicates Microglia and Immune Cells. bioRxiv 1–24 (2020).

11. Patel, T. et al. Whole-exome sequencing of the BDR cohort: evidence to support the role of the PILRA gene in Alzheimer’s disease. Neuropathol. Appl. Neurobiol. 44, 506–521 (2018).

12. Raghavan, N. S. et al. Whole-exome sequencing in 20,197 persons for rare variants in Alzheimer’s disease. Ann. Clin. Transl. Neurol. 1–11 (2018) doi:10.1002/acn3.582.

13. Prokopenko, D. et al. Whole-genome sequencing reveals new Alzheimer’s disease– associated rare variants in loci related to synaptic function and neuronal development. Alzheimer’s Dement. alz.12319 (2021) doi:10.1002/alz.12319.

14. Boyle, A. P. et al. Annotation of functional variation in personal genomes using RegulomeDB. Genome Res. 22, 1790–1797 (2012).

15. Allen, M. et al. Late-onset Alzheimer disease risk variants mark brain regulatory loci. Neurol. Genet. 1, e15 (2015).

16. Karch, C. M. et al. Alzheimer’s Disease Risk Polymorphisms Regulate Gene Expression in the ZCWPW1 and the CELF1 Loci. PLoS One 11, e0148717 (2016).

17. Aguet, F. et al. The GTEx Consortium atlas of genetic regulatory effects across human tissues. Science (80-.). 369, 1318–1330 (2020).

18. Ng, B. et al. An xQTL map integrates the genetic architecture of the human brain’s transcriptome and epigenome. Nat. Neurosci. 1–15 (2017) doi:10.1038/nn.4632.

19. Võsa, U. et al. Unraveling the polygenic architecture of complex traits using blood eQTL meta-analysis. bioRxiv 1–57 (2018).

20. Kerimov, N. et al. eQTL catalogue: A compendium of uniformly processed human gene expression and splicing QTLs. bioRxiv (2020) doi:10.1101/2020.01.29.924266.

21. Kikuchi, M. et al. Enhancer variants associated with Alzheimer’s disease affect gene expression via chromatin looping. BMC Med. Genomics 12, 128 (2019).

22. Rathore, N. et al. Paired Immunoglobulin-like Type 2 Receptor Alpha G78R variant alters ligand binding and confers protection to Alzheimer’s disease. PLoS Genet. 14, e1007427 (2018).

23. Mousseau, D. D., Banville, D., L’Abbé, D., Bouchard, P. & Shen, S. H. PILRα, a novel immunoreceptor tyrosine-based inhibitory motif-bearing protein, recruits SHP-1 upon tyrosine phosphorylation and is paired with the truncated counterpart PILRβ. J. Biol. Chem. 275, 4467–4474 (2000).

24. He, F. et al. Structural insight into the zinc finger CW domain as a histone modification reader. Structure 18, 1127–1139 (2010).

25. Li, M. et al. The histone modification reader ZCWPW1 is required for meiosis prophase I in male but not in female mice. Sci. Adv. 5, (2019).

26. Mahgoub, M. et al. Dual histone methyl reader ZCWPW1 facilitates repair of meiotic double strand breaks in male mice. Elife 9, 1–25 (2020).

27. Wells, D. et al. ZCWPW1 is recruited to recombination hotspots by PRDM9 and is essential for meiotic double strand break repair. Elife 9, 1–36 (2020).

28. Huang, T. et al. The histone modification reader ZCWPW1 links histone methylation to PRDM9-induced double-strand break repair. Elife 9, 1–48 (2020).

29. McKhann, G. et al. Clinical diagnosis of alzheimer’s disease: Report of the NINCDS-ADRDA work group⋆ under the auspices of department of health and human services task force on alzheimer’s disease. Neurology 34, 939–944 (1984).

30. McKhann, G. M. et al. The diagnosis of dementia due to Alzheimer’s disease: Recommendations from the National Institute on Aging-Alzheimer’s Association workgroups on diagnostic guidelines for Alzheimer’s disease. Alzheimer’s Dement. 7, 263–269 (2011).

31. Hyman, B. T. et al. National Institute on Aging–Alzheimer’s Association guidelines for the neuropathologic assessment of Alzheimer’s disease. Alzheimer’s Dement. 8, 1– 13 (2012).

32. Nasreddine, Z. et al. The Montreal Cognitive Assessment, MoCA: a brief screening tool for mild cognitive impairment. J. Am. Geriatr. Soc. 53, 695–699 (2005).

33. Weckx, S. et al. novoSNP, a novel computational tool for sequence variation discovery. Genome Res. 15, 436–442 (2005).

34. Li, H. & Durbin, R. Fast and accurate long-read alignment with Burrows-Wheeler transform. Bioinformatics 26, 589–595 (2010).

35. Li, H. et al. The Sequence Alignment/Map format and SAMtools. Bioinformatics 25, 2078–2079 (2009).

36. Krzywinski, M. et al. Circos: an information aesthetic for comparative genomics. Genome Res. 19, 1639–45 (2009).

37. McKenna, A. et al. The Genome Analysis Toolkit: A MapReduce framework for analyzing next-generation DNA sequencing data. Genome Res. 20, 1297–1303 (2010).

38. Depristo, M. A. et al. A framework for variation discovery and genotyping using next-generation DNA sequencing data. Nat. Genet. 43, 491–501 (2011).

39. Van der Auwera, G. A. et al. From fastQ data to high-confidence variant calls: The genome analysis toolkit best practices pipeline. Current Protocols in Bioinformatics (2013). doi:10.1002/0471250953.bi1110s43.

40. Danecek, P. et al. The variant call format and VCFtools. Bioinformatics 27, 2156–2158 (2011).

41. Lindenbaum, P. & Redon, R. Bioalcidae, samjs and vcffilterjs: Object-oriented formatters and filters for bioinformatics files. Bioinformatics 34, 1224–1225 (2018).

42. Wang, K., Li, M. & Hakonarson, H. ANNOVAR: Functional annotation of genetic variants from high-throughput sequencing data. Nucleic Acids Res. 38, 1–7 (2010).

43. Cingolani, P. et al. A program for annotating and predicting the effects of single nucleotide polymorphisms, SnpEff. Fly (Austin). 6, 80–92 (2012).

44. Rentzsch, P., Witten, D., Cooper, G. M., Shendure, J. & Kircher, M. CADD: predicting the deleteriousness of variants throughout the human genome. Nucleic Acids Res. 29, 225–229 (2018).

45. Karczewski, K. J. et al. The mutational constraint spectrum quantified from variation in 141,456 humans. Nature 581, 434–443 (2020).

46. Lek, M. et al. Analysis of protein-coding genetic variation in 60,706 humans. Nature 536, 285–291 (2016).

47. Guerreiro, R. et al. A comprehensive assessment of benign genetic variability for neurodegenerative disorders. bioRxiv (2018).

48. Price, A. L. et al. Long-Range LD Can Confound Genome Scans in Admixed Populations. Am. J. Hum. Genet. 83, 132–135 (2008).

49. Auton, A. et al. A global reference for human genetic variation. Nature 526, 68–74 (2015).

50. Chang, C. C. et al. Second-generation PLINK: Rising to the challenge of larger and richer datasets. Gigascience 4, 1–16 (2015).

51. Lee, S. et al. Optimal unified approach for rare-variant association testing with application to small-sample case-control whole-exome sequencing studies. Am. J. Hum. Genet. 91, 224–37 (2012).

52. Broman, K. W., Murray, J. C., Sheffield, V. C., White, R. L. & Weber, J. L. Comprehensive human genetic maps: Individual and sex-specific variation in recombination. Am. J. Hum. Genet. 63, 861–869 (1998).

53. Untergasser, A. et al. Primer3-new capabilities and interfaces. Nucleic Acids Res. 40, 1–12 (2012).

54. De Roeck, A. et al. Deleterious ABCA7 mutations and transcript rescue mechanisms in early onset Alzheimer’s disease. Acta Neuropathol. 134, 475–487 (2017).

55. Li, H. Minimap2: Pairwise alignment for nucleotide sequences. Bioinformatics 34, 3094–3100 (2018).

56. Thorvaldsdóttir, H., Robinson, J. T. & Mesirov, J. P. Integrative Genomics Viewer (IGV): High-performance genomics data visualization and exploration. Brief. Bioinform. 14, 178–192 (2013).

57. Martin, M. et al. WhatsHap : fast and accurate read-based phasing. bioRxiv 1–18 (2016) doi:http://dx.doi.org/10.1101/085050.

58. Cruts, M. et al. Null mutations in progranulin cause ubiquitin-positive frontotemporal dementia linked to chromosome 17q21. Nature 442, 920–924 (2006).

59. Puoti, G. et al. A mutation in the 5′-UTR of GRN gene associated with frontotemporal lobar degeneration: Phenotypic variability and possible pathogenetic mechanisms. J. Alzheimer’s Dis. 42, 939–947 (2014).

60. Liu, X., Jiao, B. & Shen, L. The Epigenetics of Alzheimer’s Disease: Factors and Therapeutic Implications. Front. Genet. 9, 1–10 (2018).

61. Fairfax, B. P. et al. Innate immune activity conditions the effect of regulatory variants upon monocyte gene expression. Science (80-.). 343, (2014).

62. Del-Aguila, J. L. et al. A single-nuclei RNA sequencing study of Mendelian and sporadic AD in the human brain. Alzheimers. Res. Ther. 11, 71 (2019).

