## Supplementary Information for "*ZCWPW1* loss-of-function variants in Alzheimer’s Disease"

[$] The following members of the BELNEU Consortium have contributed to the clinical and pathological phenotyping and follow-up of the Belgian patient cohorts: Johan Goeman, Roeland Crols (Hospital Network Antwerp (ZNA) Middelheim and Hoge Beuken, Antwerp, Belgium); Dirk Nuytten (Hospital Network Antwerp (ZNA) Stuivenberg, Antwerp, Belgium); Rudy Mercelis (Antwerp University Hospital, Edegem); Mathieu Vandenbulcke (University of Leuven and University Hospitals Leuven, Leuven, Belgium); Anne Sieben, Jan L. De Bleecker, Patrick Santens (University Hospital Ghent, Ghent, Belgium); Jan Versijpt, Alex Michotte (University Hospital Brussels, Brussels, Belgium); Olivier Deryck, Ludo Vanopdenbosch, Bruno Bergmans (AZ Sint-Jan Brugge, Bruges, Belgium); Christiana Willems, Nina De Klippel (Jessa Hospital, Hasselt, Belgium); Jean Delbeck (General Hospital Sint-Maria, Halle); Adrian Ivanoiu (Saint-Luc University Hospital, Université Catholique de Louvain, Louvain-la-Neuve, Belgium); and Eric Salmon (University of Liege and Memory Clinic, CHU Liege, Liege, Belgium).

*Corresponding author: Prof. Dr. Kristel Sleegers MD PhD

Complex Genetics of Alzheimer's Disease Group; VIB Center for Molecular Neurology

University of Antwerp - CDE

Universiteitsplein 1, B-2610, Antwerp, Belgium

### Supplementary Figures

**Supplementary Figure 1.** Genetic ancestry analysis of the case-control cohort with the 1KG samples. The PCA plot was generated on first and second genetic principal components calculated from genotyping array data of *n*=1803 samples that were included in this study, together with 1KG Project samples (*n*=2504). AFR: African population, AMR: Ad mixed American population, EAS: East Asian population, EUR: European population, SAS: South Asian population.

**Supplementary Figure 2.** Coverage Circos plot for targeted exons and UTRs of *PILRB*, *PILRA* and *ZCWPW1*. *PILRB*, *PILRA*, and *ZCWPW1* exons are colored in blue, red, and green respectively. Each exon is extended by 20 bp to include splice regions. The numbers in the inner rim indicate the exon number of each gene of interest. The outer rim indicates percentage of the study cohort covered ≥20X on the corresponding position and indicate 10% difference in this scale. The coverage information indicated on the figure is solely based on sequencing on the MiSeq platform, therefore does not include additional Sanger sequencing of *PILRB* exon 5 and *ZCWPW1* exon 14 due to relative drop in ≥20X coverage in the study cohort.

**Supplementary Figure 3.** Haplotype sharing analysis of non-singleton *ZCWPW1* predicted LoF variant carriers. **(A)** Haplotype sharing analysis for *ZCWPW1* c.631+1G>T. **(B)** Haplotype sharing analysis for *ZCWPW1* c.-29-1G>A. For each panel, the distances to the respective mutation are provided. Allele frequencies for each STR genotype are given in base lengths. Frequencies of indicated STR lengths are shown in brackets (calculated based on STR lenghts in *n*=172 Flanders-Belgian healthy control individuals) and for SNPs gnomAD NFE MAFs are provided. IDs of STRs and SNPs within the targeted resequencing locus are bolded. "?" indicates that the related SNP genotype information is unknown. Risk-increasing and mutation alleles are bolded in red. Main possible shared haplotypes are highlighted in green and blue respectively.

**Supplementary Figure 4.** IGV snapshot of Illumina short-read sequencing reads to show phasing between *ZCWPW1* p.Glu105Gly and *ZCWPW1* p.Glu95Lys predicted deleterious rare missense variants in LOAD-P7.

**Supplementary Figure 5.** IGV snapshot of Nanopore long-read sequencing reads of LOAD-P2 and LOAD-P3 harboring rs34919929 and rs774275324 (*ZCWPW1* c.631+1G>T). Aligned reads are colored by their "HP" tag (haplotype identifier) added by WhatsHap software for each sample separately. For each sample, haplotypes defined by WhatsHap are shown below its coverage track. The coverage tracks are in $\log_{10}$ scale.

**Supplementary Figure 6.** IGV snapshot of Nanopore long-read sequencing reads of LOAD-P4, LOAD-P5 and LOAD-P6 harboring rs34919929 and rs1180932049 (*ZCWPW1* c.-29-1G>A). Aligned reads are colored by their "HP" tag (haplotype identifier) added by WhatsHap software for each sample separately. For each sample, haplotypes defined by WhatsHap shown below its coverage track. The coverage tracks are in $\log_{10}$ scale.

**Supplementary Figure 7.** IGV snapshot of Nanopore long-read sequencing reads of three negative control samples having all possible genotypes for rs34919929 and not harboring any predicted LoF mutations of interest. The negative control samples were named according to their rs34919929 genotype. Aligned reads are colored by their "HP" tag (haplotype identifier) added by WhatsHap software for each sample separately. For each sample, haplotypes defined by WhatsHap shown below its coverage track. The coverage tracks are in $\log_{10}$ scale.

**Supplementary Figure 1.** Genetic ancestry analysis of the case-control cohort with the 1KG samples.

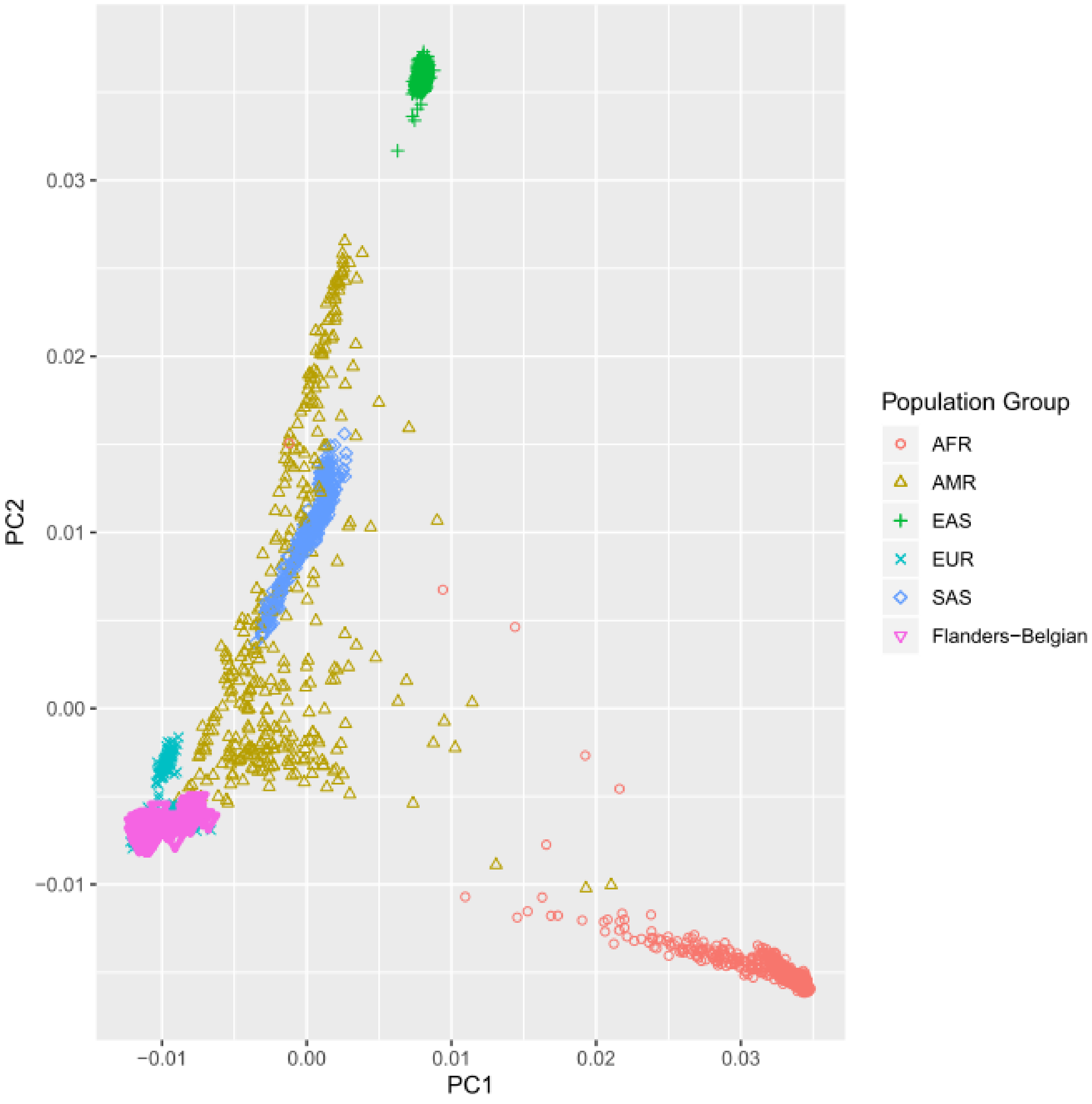

**Supplementary Figure 2.** Coverage Circos plot for targeted exons and UTRs of *PILRB*, *PILRA* and *ZCWPW1*.

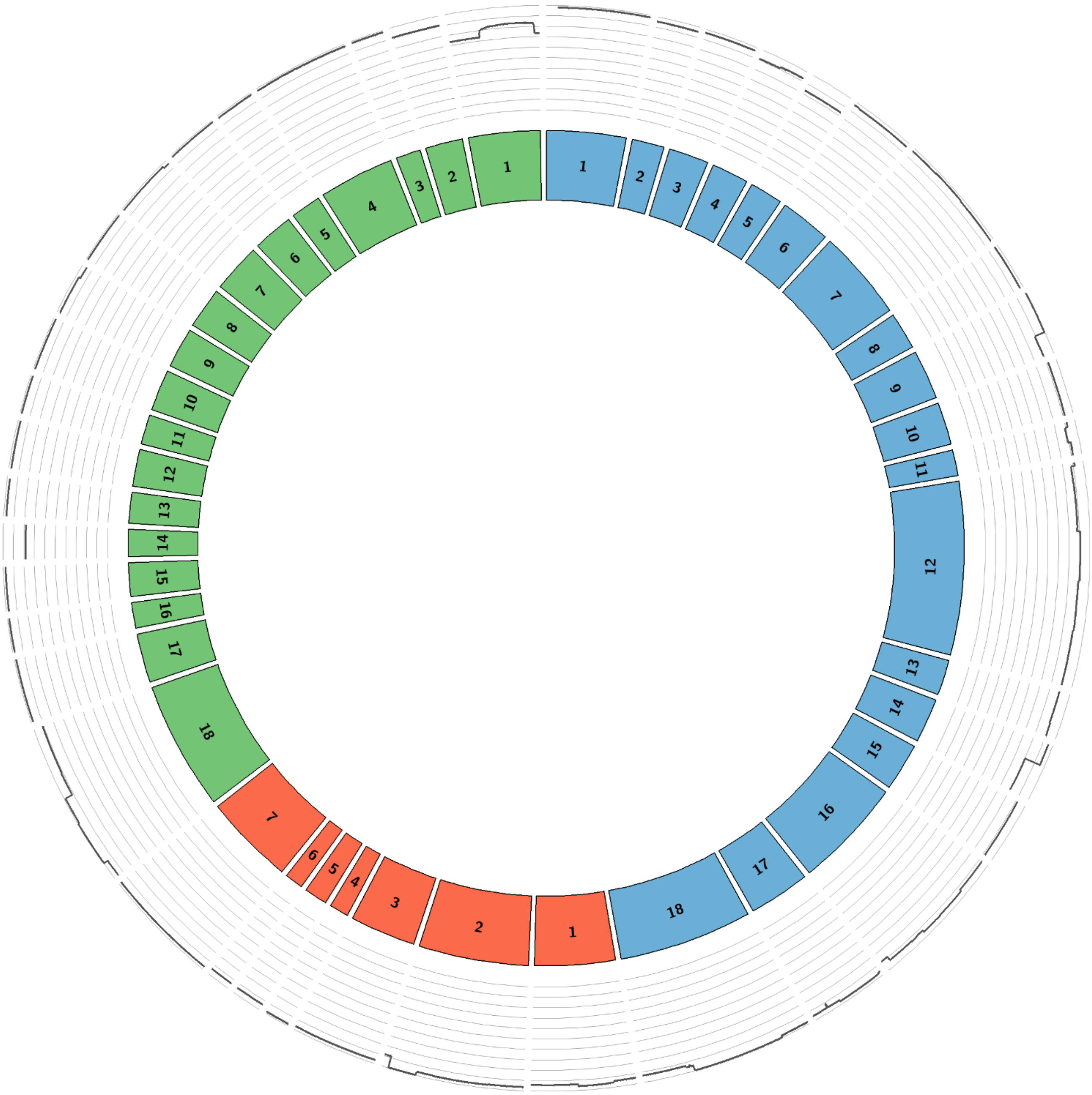

**Supplementary Figure 3.** Haplotype sharing analysis of non-singleton *ZCWPW1* predicted LoF variant carriers. **(A)** Haplotype sharing analysis for *ZCWPW1* c.631+1G>T. **(B)** Haplotype sharing analysis for *ZCWPW1* c.-29-1G>A.

## A.

| | | ***ZCWPW1* c.631+1G>T** | | | | |
|---|---|---|---|---|---|---|
| | | **LOAD-P2** | | **LOAD-P3** | | |
| **Distance to LoF** | **STR or *SNP*** | **Allele 1** | **Allele 2** | **Allele 1** | **Allele 2** | **Common STR Allele Frequencies or *gnomAD NFE MAF*** |
| -4.87 Mb | D7S2431 | 116 | 118 | 102 | 114 | - |
| -4.39 Mb | *rs1488514* | A | A | A | A | *G (23.3%)* |
| -3.46 Mb | D7S1796 | 287 | 292 | 287 | 284 | 287 (28.4%) |
| -2.68 Mb | D7S554 | 260 | 254 | 260 | 262 | 260 (33.7%) |
| -2.09 Mb | *rs6970990* | T | T | G | G | *G (47.7%)* |
| -1.46 Mb | D7S651 | **178** | 182 | **178** | 178 | 178 (36.6%) |
| -745 Kb | D7S647 | **178** | 180 | **178** | 178 | 178 (38.6%) |
| - 9.48 Kb | ***rs1476679*** | **T** | C | **T** | C | *C (29.7%)* |
| -1.59 Kb | ***rs34919929*** | **A** | G | **A** | G | *G (29.7%)* |
| 0 | ***rs774275324 (c.631+1G>T)*** | **A** | C | **A** | C | *A (0.0023%)* |
| +42 Kb | D7S2480 | **213** | 223 | **213** | 223 | 213 (8.4%) & 223 (1.4%) |
| +763 Kb | D7S477 | **224** | 226 | **224** | 224 | 224 (37.2%) |
| +1.73 Mb | D7S666 | **161** | 165 | **161** | 161 | 161 (43.8%) |
| +2.09 Mb | D7S2448 | **249** | 237 | **249** | 251 | 249 (29.9%) |
| +3.31 Mb | *rs727708* | T | T | C | C | *T (48.5%)* |
| +3.45 Mb | D7S2504 | 200 | 208 | 194 | 212 | - |
| +3.53 Mb | *rs10245317* | G | A | A | G | *G (16.6%)* |
| +4.03 Mb | D7S2494 | 286 | 292 | 292 | 296 | 292 (6.1%) |

**Supplementary Figure 3 continued**

## B.

| | | *ZCWPW1* c.-29-1G>A | | | | | | |
|---|---|---|---|---|---|---|---|---|
| | | LOAD-P4 | | LOAD-P5 | | LOAD-P6 | | |
| **Distance to LoF** | **STR or *SNP*** | **Allele 1** | **Allele 2** | **Allele 1** | **Allele 2** | **Allele 1** | **Alelle 2** | **Common STR Allele Frequencies or *gnomAD NFE MAF*** |
| -4.87 Mb | D7S2431 | 102 | 118 | 114 | 122 | 112 | 118 | 118 (17.7%) |
| -4.39 Mb | *rs1488514* | G | G | ? | ? | A | A | *G (23.3%)* |
| -3.46 Mb | D7S1796 | **287** | 296 | **287** | 296 | **287** | 287 | 287 (28.4%) & 296 (15.4%) |
| -2.69 Mb | D7S554 | **254** | 260 | **254** | 254 | **254** | 258 | 254 (34%) |
| -2.09 Mb | *rs6970990* | **T** | G | **?** | ? | **T** | T | *G (47.7%)* |
| -1.46 Mb | D7S651 | **182** | 178 | **182** | 182 | **182** | 182 | 182 (28.4%) |
| -749 Kb | D7S647 | **178** | 176 | **178** | 178 | **178** | 180 | 178 (38.6%) |
| -13.85 Kb | ***rs1476679*** | **T** | C | **T** | T | **T** | C | *C (29.7%)* |
| -5.96 Kb | ***rs34919929*** | **A** | G | **A** | A | **A** | G | *G (29.7%)* |
| 0 | ***rs1180932049 (c.-29-1G>A)*** | **T** | C | **T** | C | **T** | C | *T (0.0023%)* |
| +37 Kb | D7S2480 | **202** | 221 | **202** | 217 | **202** | 221 | 202 (21.2%) & 221 (20%) |
| +758 Kb | D7S477 | **226** | 226 | **226** | 226 | **226** | 224 | 226 (18.8%) |
| +1.73 Mb | D7S666 | **159** | 165 | 165 | 161 | **159** | 161 | 159 (21.2%) & 161 (43.8%) & 165 (11.9%) |
| +2.08 Mb | D7S2448 | **247** | 249 | 247 | 243 | **247** | 249 | 247 (13%) |
| +3.31 Mb | *rs727708* | **T** | C | ? | ? | **T** | C | *T (48.5%)* |
| +3.44 Mb | D7S2504 | **194** | 204 | 194 | 206 | **194** | 210 | 194 (12.7%) |
| +3.53 Mb | *rs10245317* | A | A | ? | ? | G | G | *G (16.6%)* |
| +4.03 Mb | D7S2494 | 292 | 286 | 294 | 298 | 292 | 296 | 292 (6.1%) |

**Supplementary Figure 4.** IGV snapshot of Illumina short-read sequencing reads to show phasing between *ZCWPW1* p.Glu105Gly and *ZCWPW1* p.Glu95Lys predicted deleterious rare missense variants in LOAD-P7.

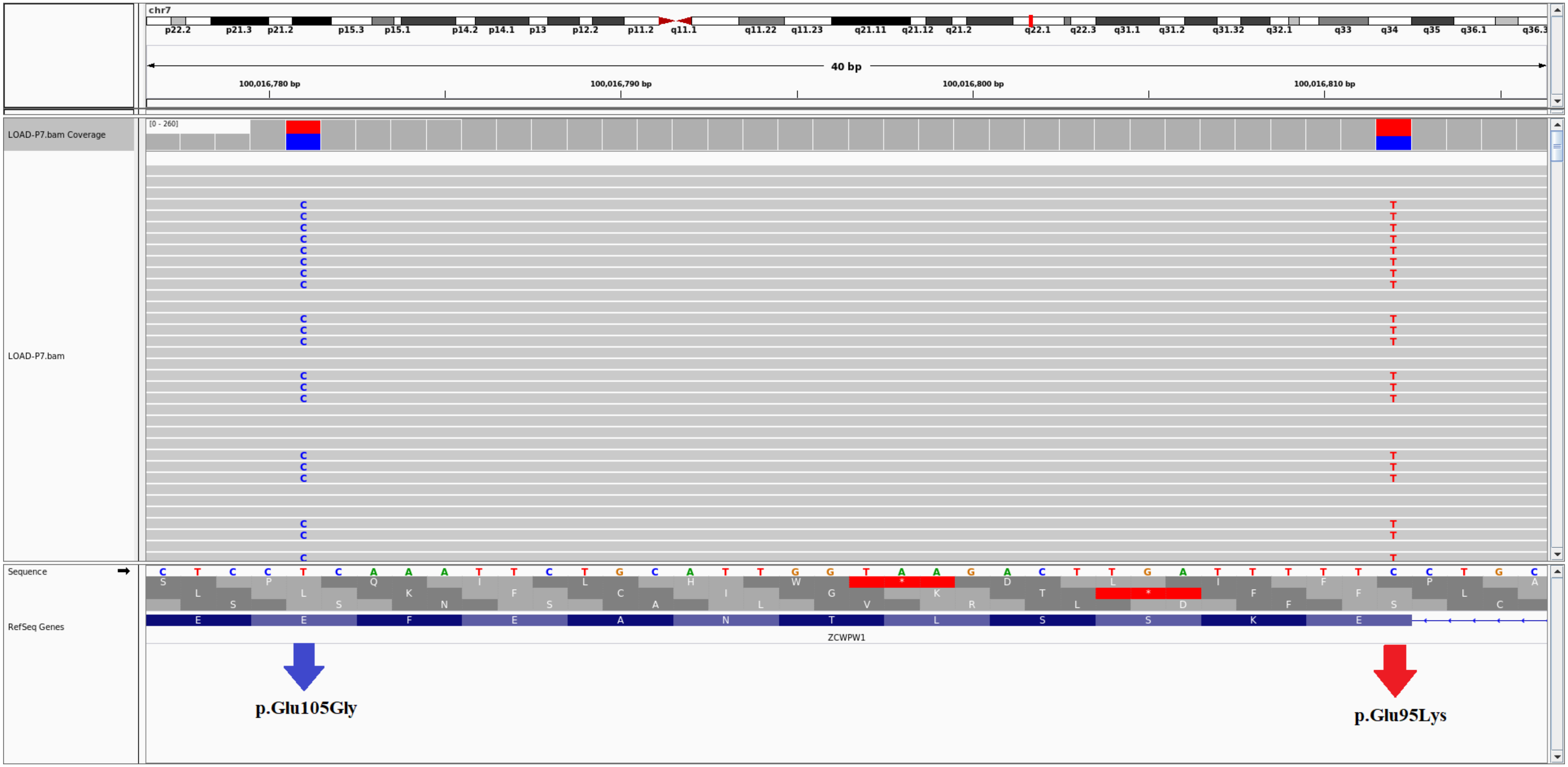

**Supplementary Figure 5.** IGV snapshot of Nanopore long-read sequencing reads of LOAD-P2 and LOAD-P3 harboring rs34919929 and rs774275324 (*ZCWPW1* c.631+1G>T).

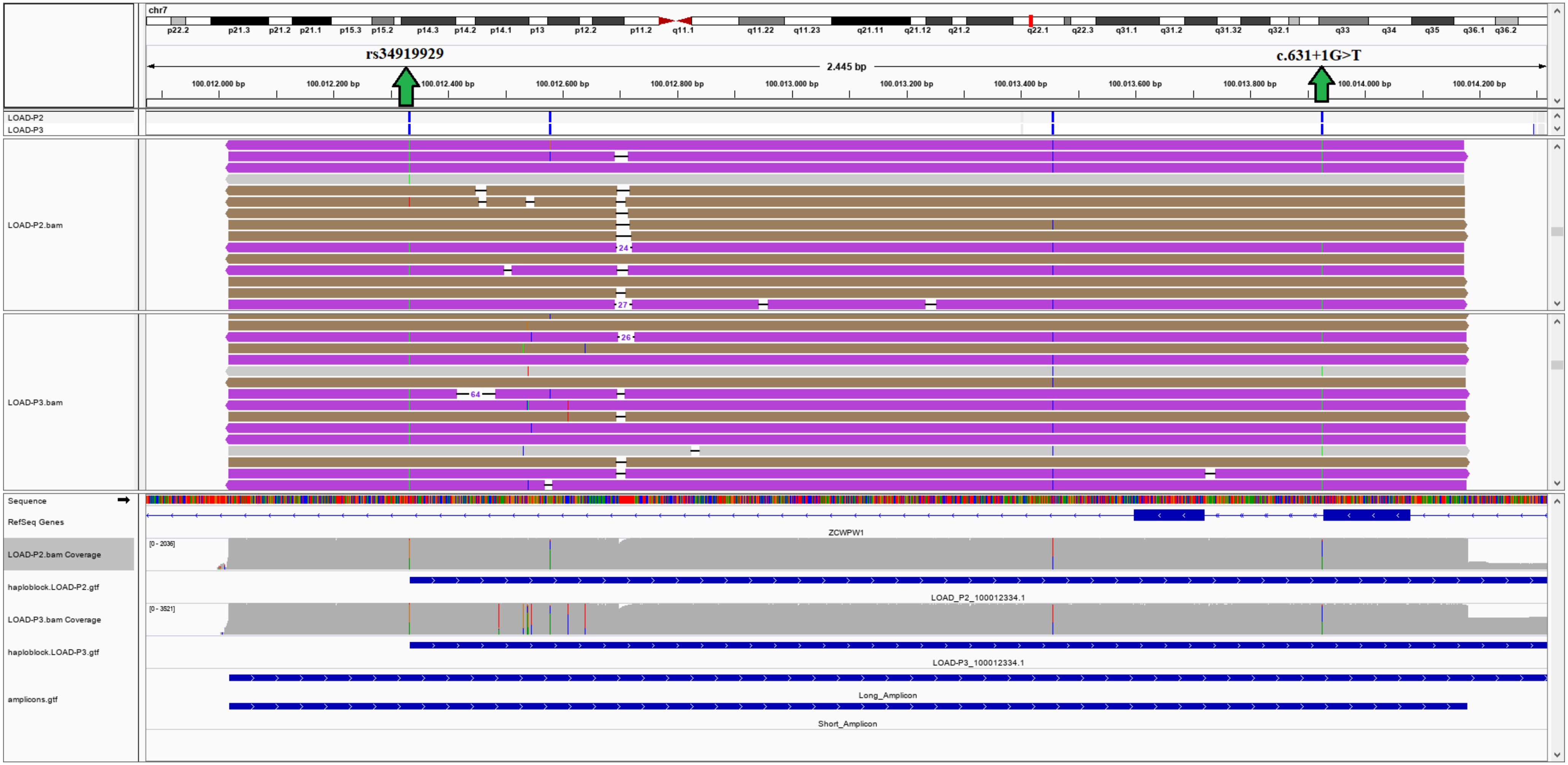

**Supplementary Figure 6.** IGV snapshot of Nanopore long-read sequencing reads of LOAD-P4, LOAD-P5 and LOAD-P6 harboring rs34919929 and rs1180932049 (*ZCWPW1* c.-29-1G>A).

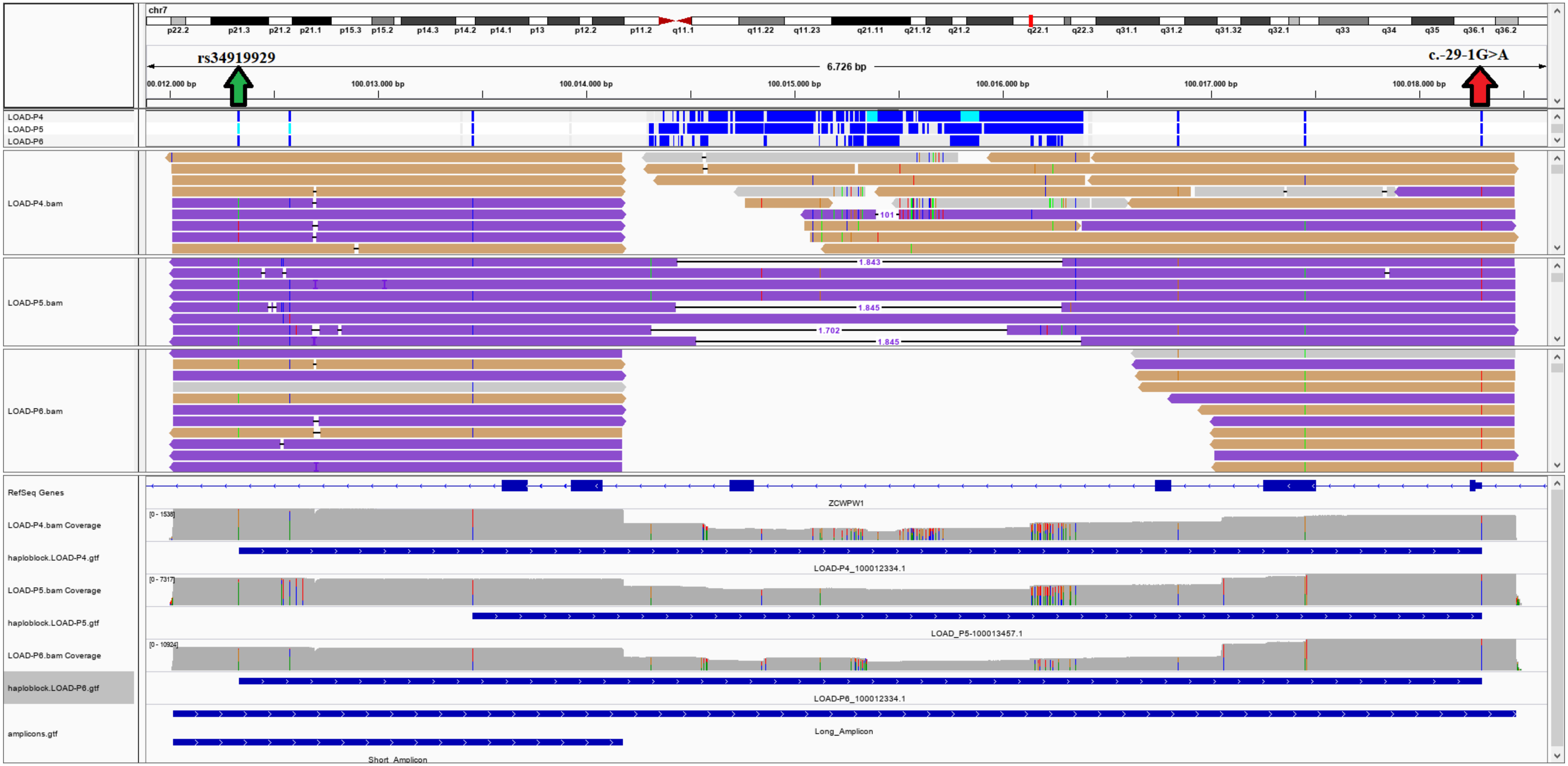

**Supplementary Figure 7.** IGV snapshot of Nanopore long-read sequencing reads of three negative control samples having all possible genotypes for rs34919929 and not harboring any predicted LoF mutations of interest.

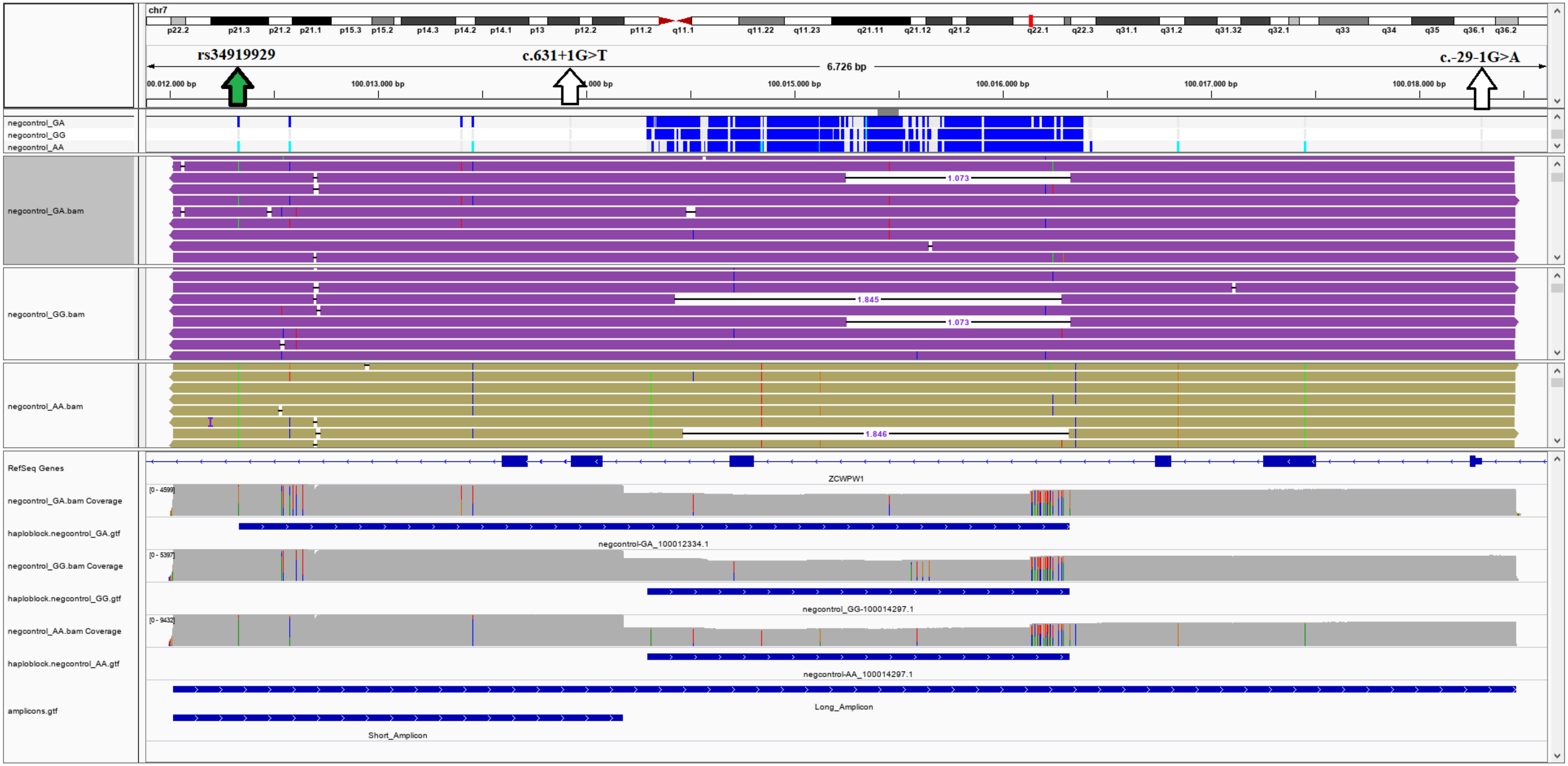
